# Beyond Seizure Burden: Seizure Semiology, but not Frequency, is Associated with Caregiver-Reported Autistic Behaviors in SYNGAP1-DEE

**DOI:** 10.64898/2026.04.19.26351217

**Authors:** L. Kiwull, V. Schmeder, M. Zenker, M. Mengual Hinojosa, J.R. Perkins, J. Ranea, G. Kluger, T. Hartlieb, M. Pringsheim, C. von Stülpnagel, D. Weghuber, K. Eschermann

**Author notes:** **Corresponding Author: Kirsten Eschermann**.

## Abstract

1.

**Purpose:** SYNGAP1-related developmental and epileptic encephalopathy (SYNGAP1-DEE) is characterized by high rates of both epilepsy and autism spectrum disorder (ASD). While the clinical spectrum is well-documented, the link between specific seizure semiologies and caregiver-reported autistic behaviors is not well understood. This study analyzed the correlation between ten distinct seizure types, their frequencies, and a caregiver-reported autistic behavior score.

**Method:** Clinical data were extracted from the PATRE (PATient-based phenotyping and evaluation of therapy for Rare Epilepsies) Registry for SYNGAP1, in the framework of the EURAS project (Grant No. 101080580, Horizon Europe). This study employed a retrospective cross-sectional analysis of caregiver-reported registry data. Analysis was restricted to an analytic cohort of N=337 participants with complete data for both the epilepsy questionnaire (including epilepsy status, seizure semiology, and peak seizure frequency items) and the behavior questionnaire (from a total N=522 registry participants). Caregiver-reported autistic behaviors were quantified using a standardized caregiver-reported scale (Likert 1–5). Statistical associations were evaluated using the Wilcoxon rank-sum test to compare caregiver-reported autistic behavior scores across different seizure semiologies and Spearman’s rank correlation to assess the impact of seizure frequency (9-point scale).

**Results:** Within the analytic cohort (N=337), epilepsy was reported in 259 patients. Eyelid myoclonia was the most prevalent semiology, affecting 64.9% (n=168) of the epilepsy-positive group. Atypical absences (n=77) demonstrated the most profound and statistically robust association with higher caregiver-reported autistic behavior scores (FDR-adjusted p = 0.001). Significant associations were also observed for typical absences (n=70, FDR-adjusted p = 0.018), eyelid myoclonia (FDR-adjusted p = 0.018), myoclonic-atonic seizures (n=40, FDR-adjusted p = 0.019), and atonic seizures (n=72, FDR-adjusted p = 0.025). Focal and tonic-clonic seizures showed weaker associations (FDR-adjusted p = 0.026 and p = 0.047, respectively).

Crucially, quantitative analysis revealed no significant correlation between ordinal caregiver-reported peak seizure frequency ratings and caregiver-reported autistic behavior scores across all semiologies (e.g., Eyelid Myoclonia: p=0.096; Atypical Absences: p=0.744), indicating no detectable association between peak-frequency ratings and caregiver-reported autistic behavior scores.

**Conclusion:** Higher caregiver-reported autistic behavior scores in SYNGAP1-DEE were most strongly associated with the presence of atypical absences, representing a generalized, thalamocortical seizure network dysfunction. In contrast, no detectable association was observed between caregiver-reported autistic behavior scores and the ordinal caregiver-reported peak seizure frequency metric. Atypical absences and related semiologies may serve as clinical “red flags” for increased neurodevelopmental comorbidity severity, regardless of reported peak seizure frequency.

**Abstract Summary:** This study investigates the relationship between ten seizure semiologies, seizure frequency, and severity of caregiver-reported autistic behaviors in a large-scale international cohort of N=337 patients with SYNGAP1-DEE. We identify a robust association between elevated caregiverreported autistic behavior scores and specific thalamocortical seizure patterns, most prominently atypical absences.

Notably, our analysis reveals that this association is independent of seizure frequency, demonstrating no detectable association between this ordinal, caregiver-reported seizure frequency metric and caregiver-reported autistic behaviors.

## 2. Introduction

SYNGAP1-related developmental and epileptic encephalopathy (SYNGAP1-DEE) is a prototypical synaptopathy characterized by a profound clinical burden^1^, most notably the co-occurrence of treatment-resistant epilepsy and autism spectrum disorder (ASD). Since the initial description of pathogenic variants in 2009^2^, the role of the SYNGAP1 protein in regulating synaptic plasticity and excitatory-inhibitory balance has been well-established^3^, yet the clinical interplay between specific seizure phenotypes and the severity of neurodevelopmental comorbidities remains inadequately defined.

A significant gap in current clinical research is the tendency to treat epilepsy as a binary variable (presence vs. absence) or a purely quantitative measure (seizure frequency). This approach fails to account for the potential impact of specific seizure semiologies on distinct neurodevelopmental trajectories. Evidence from other developmental and epileptic encephalopathies (DEEs) suggests that different seizure types may reflect diverse underlying network dysfunctions^4^, yet large-scale data differentiating these impacts on caregiver-reported autistic behaviors are lacking. This study aims to provide large-scale empirical evidence for the observation that developmental slowing “does not always occur in concert with… frequent seizures” (Scheffer et al., p. 1017)^5^, a phenomenon that remains poorly quantified in genetic DEEs.

To address these challenges, we utilized the Patient-based Phenotyping and Evaluation of Therapy for Rare Epilepsies (PATRE) SYNGAP1 Registry. PATRE is a digital, caregiver-centered framework designed to facilitate deep phenotyping and real-time data acquisition directly from families^6^. This methodology enables the collection of high-resolution phenotypic data that traditional clinical registries often miss. Our study is integrated into the EURAS consortium (European RASopathy Network, Grant No. 101080580)^7^, an international effort aimed at stratifying patient cohorts and identifying “disease signatures” to pave the way for targeted therapeutic interventions in the neurodevelopmental RASopathies Costello Syndrome (CS), Cardiofaciocutaneous Syndrome (CFC), SYNGAP1 Syndrome and Noonan Syndrome.

In the present study, we analyzed a large international cohort of 337 patients from the PATRE registry. We hypothesized that the severity of the social-communicative phenotype in SYNGAP1-DEE is not uniformly affected by all seizure types. Specifically, we postulated that atypical absences serve as the primary clinical manifestation of a generalized, thalamocortical-mediated seizure network that is inherently more strongly associated with caregiver-reported autistic behaviors than focal seizure patterns, suggesting a shared network-level vulnerability.

## 3. Methods

### Data Source and Registry Framework

The data for this study were obtained from the **PATRE** (PATient-Based phenotyping and evaluation of therapy for Rare Epilepsies) SYNGAP1 registry. This registry is a patient-centered platform designed to collect comprehensive health data as a foundation for preclinical models in EURAS, explore genotype-phenotype correlation and serve as a natural history as a basis for clinical trials. Data collection is facilitated through a multilingual mobile app^8^, which ensures global accessibility and maximizes participation from the international rare disease community. The registry was initiated by the German SYNGAP1 patient organization in collaboration with its medical advisory board and is supported by a patient board of 13 European SYNGAP1 patient organizations.

### Patient Cohort and Inclusion Criteria

Of 522 registered participants, 337 were included in the analytic cohort based on completion of both the epilepsy questionnaire (including epilepsy status, seizure semiology, and peak seizure frequency items) and the behavior questionnaire. Participants with incomplete data in either questionnaire were excluded from analyses requiring those variables. No additional clinical filtering was applied. All analyses were performed as complete-case analyses for the variables under study. Denominators may vary across analyses due to item-level missingness, particularly for composite variables.

### Clinical Metrics and Statistical Analysis

Quantitative clinical data were gathered using standardized instruments and expert-validated scales:

### Seizure Semiology

Seizure phenotypes were classified according to the **2017 ILAE (International League Against Epilepsy) operational classification of seizure types**.^9^ Caregivers identified specific seizure semiologies from a predefined list of 10 distinct types (including *Eyelid myoclonia, Atypical/Typical absences, Myoclonic-atonic seizures*, and *Focal seizures*). To ensure classification accuracy, caregivers were provided with detailed definitions and educational materials for each seizure type. For each identified seizure type, caregivers rated the maximal historical seizure frequency—reflecting the period when the seizure burden was most pronounced—on a 9-point ordinal scale (1: “Once in a lifetime”, 2: “Less than once a year”, 3: “Several times a year”, 4: “Once a month”, 5: “Several times a month”, 6: “Once a week”, 7: “Several times a week”, 8: “Once a day”, 9: “Several times per hour”). This specific metric (maximal vs. current frequency) was selected to capture the full phenotypic potential of the underlying network dysfunction (“circuitopathy”) and to capture peak severity; however, it may not represent current burden or cumulative exposure. This focus on maximal historical frequency accounts for the fact that in certain DEE semiologies, “families often stop ‘seeing’ seizures… [which] come to be regarded as part of the patient’s normal functioning”^5^. By capturing the period of highest intensity, we minimize confounding effects from age-dependent evolution or treatment.

### Behavioral Assessment

Caregiver-reported autistic behaviors were quantified using a **5-point Caregiver Likert scale**. Caregivers rated the frequency of behaviors subjectively perceived as autistic, ranging from *‘Never’* (1) to *‘All the time’* (5), in response to the standardized question: *“Does the patient exhibit abnormal behaviors that you subjectively perceive as autistic?”*. Additionally, the status of a formal autism diagnosis was recorded. To assess the validity of this subjective metric, a sensitivity analysis was conducted comparing the Likert scores of patients with a formal ASD diagnosis versus those without. Crucially, this validation was restricted to patients who had answered “Yes” to having undergone formal autism testing, ensuring that the control group represents “confirmed negatives” rather than untested individuals.

### Statistical Computing

All statistical analyses were performed using R^10^.

#### Group Comparisons

The Wilcoxon rank-sum test was employed to identify significant differences in Caregiver-reported autistic behavior scores between patients with and without specific seizure semiologies.

#### Correlation Analysis

To assess the relationship between seizure frequency and caregiver-reported autistic behavior scores, Spearman’s rank correlation (\rho) was used.

#### Significance

To safeguard against alpha-error inflation due to multiple comparisons, primary inference used a conservative unadjusted threshold (p<0.01). In addition, Benjamini–Hochberg false discovery rate (FDR)–adjusted p-values (two-sided, q<0.05) are reported as a complementary multiple-testing sensitivity analysis. Additionally, to test the “Circuitopathy” hypothesis, a composite binary variable (“Thalamocortical Phenotype”) was created, categorizing patients as positive if they exhibited any of the associated generalized semiologies. This “Thalamocortical Phenotype” was defined a priori as the presence of any of the following generalized seizure semiologies commonly considered thalamocortical: absence seizures (typical or atypical), eyelid myoclonia, atonic, or myoclonic-atonic seizures.

### Reporting Guideline (STROBE)

This manuscript is reported in accordance with the STROBE (Strengthening the Reporting of Observational Studies in Epidemiology) statement for cross-sectional studies.

### Ethics, Consent, and Data Protection

PATRE operates under an ethics and consent framework that enables caregiver-reported data collection and secondary analyses for research purposes. The present analysis used de-identified data.

Ethics committee approval: Ethik-Kommission der Bayerischen Landesärztekammer; Number 21058; Date of approval: 25.08.2021.

All caregivers provided informed consent for participation and research use of de-identified data via the PATRE platform.

### Data Availability

De-identified data underlying the findings of this study are not publicly available due to governance and privacy constraints in a rare disease registry context. Access to de-identified data may be considered upon reasonable request to the corresponding author and/or registry governance body, subject to scientific review, data use agreements, and applicable regulatory and ethical requirements.

## 4. Results

### Cohort Characteristics

The analysis included a cohort of **337 patients** with confirmed epilepsy status (yes/no) from the PATRE registry. The population showed a median age of 9 years (IQR: 6–14), with a gender distribution of 54.4% male and 45.6% female. Within this cohort, epilepsy was reported in **259 patients (76.9%)**.

### Validation of Caregiver-Reported Autism Score

A sensitivity analysis confirmed the robustness of the caregiver-reported autistic behavior score as a proxy for clinical status. Among patients who underwent formal testing, those with a confirmed ASD diagnosis (n=134) had significantly higher caregiver-reported scores (Median: 4.0) compared to those with a formally excluded diagnosis (n=37, Median: 3.0). The difference was highly significant (Wilcoxon rank-sum test, W = 1209.5, p < 0.001), supporting the caregiver score as a proxy against clinical test results.

### Association between Seizure Semiology and Caregiver-Reported Autistic Behavior Score

We analyzed the correlation between 10 distinct seizure semiologies and caregiver-reported autistic behavior scores. To account for multiple comparisons, we prespecified a conservative unadjusted threshold (p<0.01) and additionally report FDR-adjusted p-values (Benjamini– Hochberg).

### Atypical Absences and Significant Thalamocortical Phenotypes

The analysis identified a robust pattern of behavioral severity associated with generalized discharges. Atypical Absences (29.7%, n=77) demonstrated the strongest and most distinct statistical evidence for increased caregiver-reported autistic behavior scores (FDR-adjusted p = 0.001). Significant associations were also confirmed for Typical Absences (27.0%, n=70, FDR-adjusted p = 0.018) and Eyelid Myoclonia (FDR-adjusted p = 0.018). The latter was the most prevalent semiology in the cohort, affecting 64.9% (n=168) of the epilepsy-positive group. Furthermore, Myoclonic-Atonic Seizures (15.4%, n=40, FDR-adjusted p = 0.019) and Atonic Seizures (27.8%, n=72, FDR-adjusted p = 0.025) met the strict threshold for significance.

### Weaker Associations in Focal and Motor Seizures

Crucially, while other seizure types such as Focal Seizures (9.3%, n=24, FDR-adjusted p=0.026) and Tonic-Clonic Seizures (11.6%, n=30, FDR-adjusted p=0.047) showed statistically significant associations with autistic behaviors, their effect was notably weaker and failed to reach the **highly significant threshold (p ≤ 0.001) observed for the primary clinical marker—atypical absences—or the composite thalamocortical phenotype**. Furthermore, Myoclonic Seizures (n=39, FDR-adjusted p=0.140) and Epileptic Spasms (n=16, FDR-adjusted p=0.258) showed no statistically significant group differences in caregiver-reported autistic behavior scores.

### Thalamocortical Phenotype as Network Validation

To evaluate whether the profound impact of atypical absences reflects a broader underlying network dysfunction, we compared patients reporting any semiology within the a priori thalamocortical set (n = 232) to those who did not (n = 105; total valid n = 337). Patients with the Thalamocortical Phenotype had significantly higher caregiver-reported autistic behavior scores (Median 4.0 vs 3.0; Mean 3.42 vs 2.65; Wilcoxon p < 0.001), supporting the concept of a shared underlying circuitopathy.

### Role of Age

A weak positive correlation was observed between age and caregiver-reported autistic behavior score (Spearman ρ=0.18, p=0.001), indicating a modest association (explaining 3.1% of the variance).

### Quantitative Correlation Analysis

To evaluate whether a dose-response relationship exists between seizure frequency and the caregiver-reported autistic behavior scores, we performed a Spearman rank correlation analysis across all ten semiologies.

No significant correlation was observed between seizure frequency (scale 1–9) and the caregiver-reported autistic behavior score (scale 1–5) for any semiology. Even for the most prevalent semiologies associated with elevated caregiver-reported autistic behavior scores, Spearman correlations remained low and non-significant: Eyelid Myoclonia (n=169, ρ = 0.13, p = 0.096), Atypical Absences (n=78, ρ = 0.04, p = 0.744), and Typical Absences (n=71, ρ = 0.13, p = 0.282). Similarly, no correlation was found for motor seizure types, such as Tonic– Clonic seizures (n=30, ρ = 0.07, p = 0.718) or Focal seizures (n=24, ρ = -0.19, p = 0.385).

## Visualization

Figure 1. summarizes univariate group differences in caregiver-reported autistic behavior scores between participants with versus without each seizure semiology. **Figure 2** shows Spearman rank correlations between the ordinal peak-frequency ratings and caregiver-reported autistic behavior scores across semiologies.

**Figure 1.**
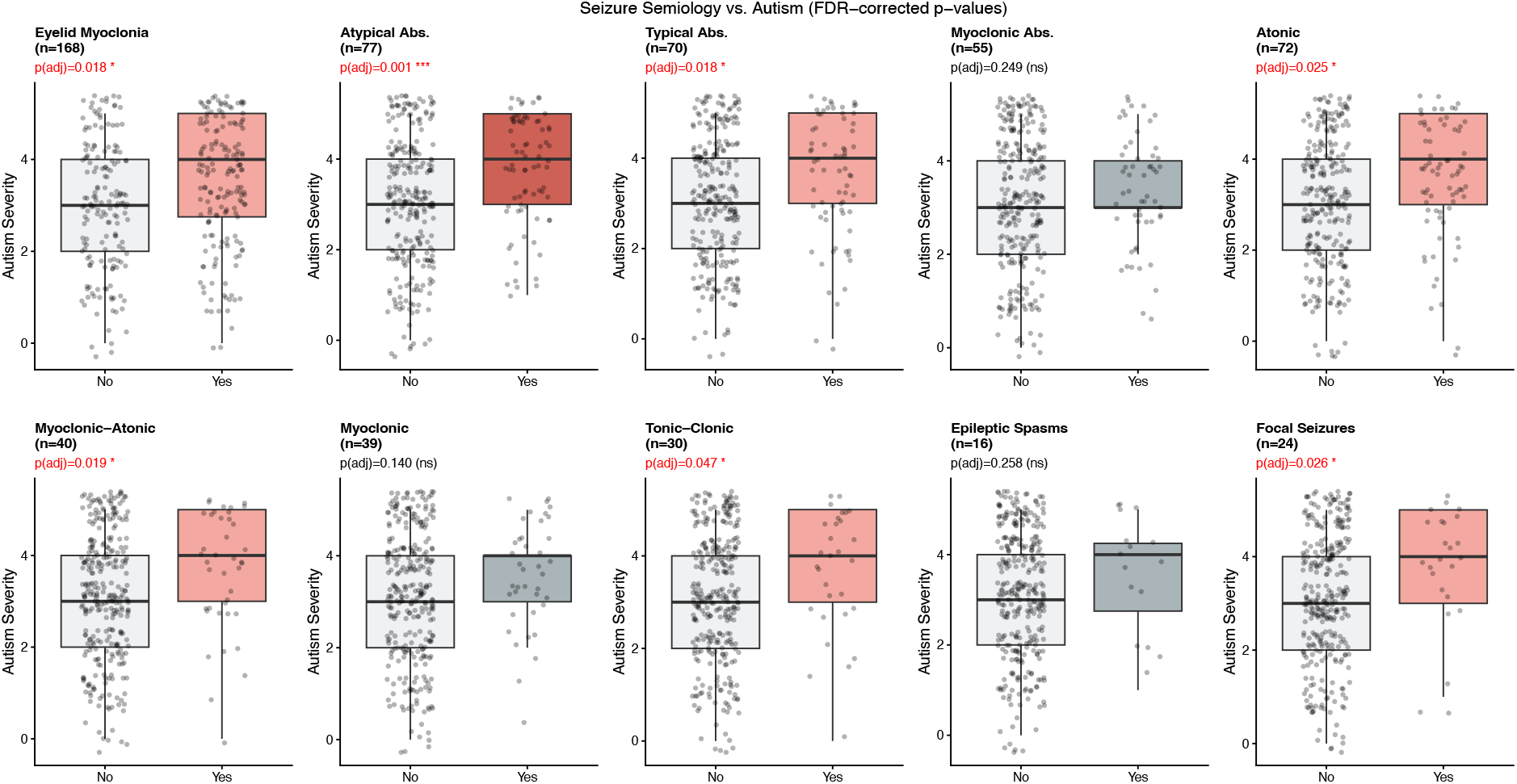
Univariate group differences in caregiver-reported autistic behavior scores (Likert 1– 5) between participants with versus without each seizure semiology (Wilcoxon rank-sum tests). Significance is reported using Benjamini-Hochberg false discovery rate (FDR)-adjusted p-values (* p < 0.05, *** p < 0.001).

**Figure 2.**
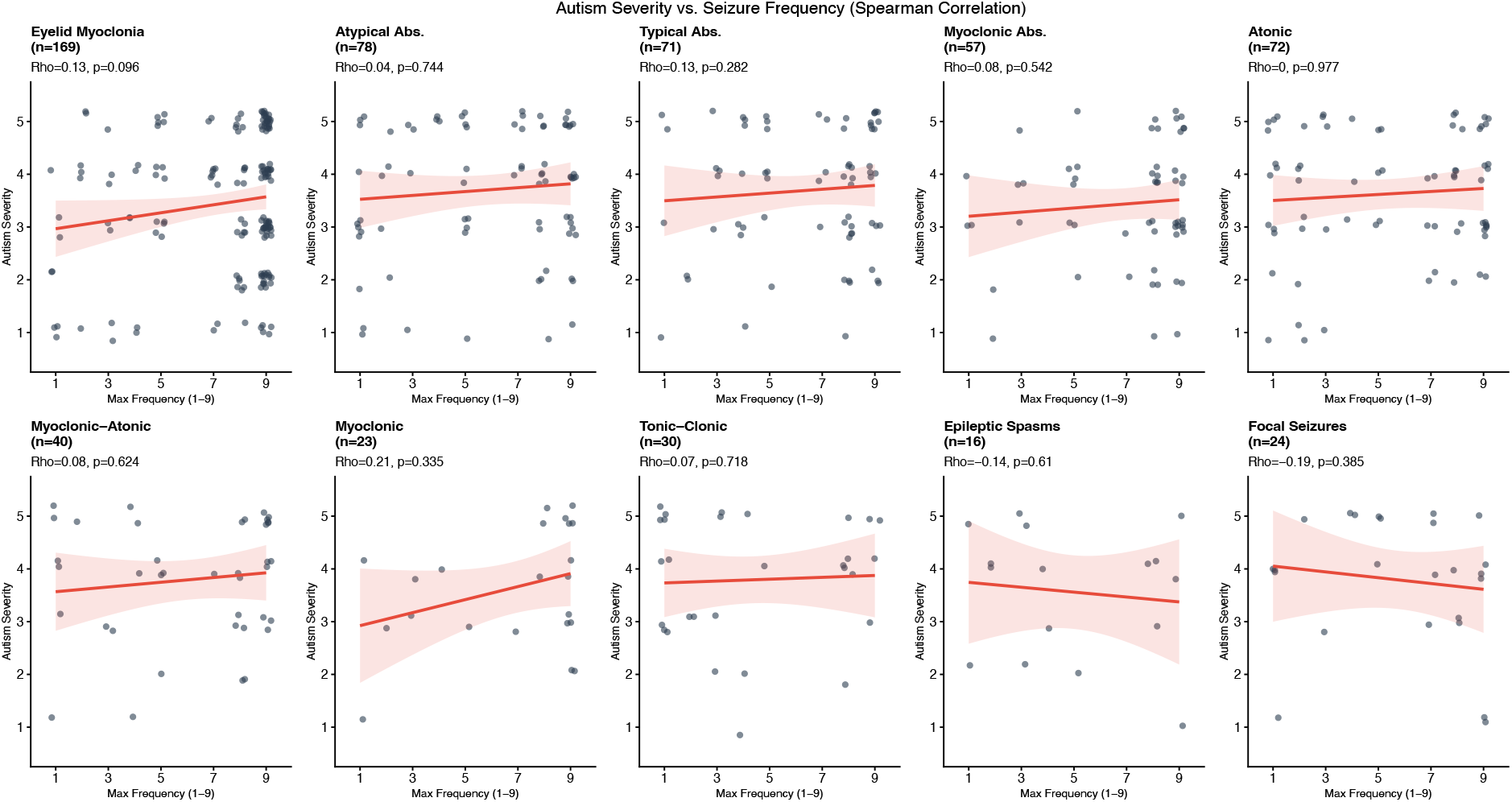
Spearman rank correlations between caregiver-reported autistic behavior scores (Likert 1–5) and ordinal caregiver-reported peak seizure frequency ratings (scale 1–9) across the ten seizure semiologies. Peak frequencies were derived from combined historical and current maximal frequency data. Each panel shows individual participants, the fitted trend line with 95% confidence band, and the Spearman ρ and p-value. Sample sizes vary by semiology because correlations are restricted to participants reporting the respective seizure type and providing a valid peak-frequency rating.

## 5. Discussion

### Interpretation of the Atypical Absence and Thalamocortical Phenotype

Our data demonstrate that the occurrence of atypical absences may serve as a clinical marker for severe caregiver-reported autistic behaviors in SYNGAP1-DEE. These behavioral impairments probably are not a consequence of general epileptic burden but may be intrinsically linked to an underlying thalamocortical network dysfunction, further supported by the high significance of the broader thalamocortical phenotype (including eyelid myoclonia and atonic seizures). The relatively weaker correlation with focal seizures (FDR-adjusted p=0.026) and generalized tonic-clonic seizures (FDR-adjusted p=0.047) supports the hypothesis that specific network dysfunctions underlie both the epilepsy and the core behavioral phenotype, with the thalamocortical network acting as the primary driver.

Crucially, our quantitative analysis revealed no detectable association between caregiver-reported seizure frequency (ordinal scale) and caregiver-reported autistic behavior scores. These findings provide a direct empirical response to the knowledge gap identified by Scheffer et al. (2025), who stated that “it is unclear if findings [seizure counts] will correlate with changes in cognition or other parameters” (p. 1022)^5^. Our data suggest that in SYNGAP1-DEE, the qualitative nature of the circuitopathy is a far more reliable indicator of severity of behavioral abnormalities than quantitative seizure counts. While the presence of thalamocortical semiologies is highly predictive of social-communicative impairment, the frequency of these seizures (ranging from rare to several times per hour) showed no significant correlation with caregiver-reported autistic behavior scores (all p > 0.30). This suggests that the neurobehavioral phenotype is driven by the qualitative nature of the underlying circuitopathy rather than a quantitative “dose-response” effect of seizure frequency as captured by this caregiver-reported ordinal measure.

We propose that this correlation is driven by shared pathophysiological mechanisms within the thalamocortical circuitry. The thalamus functions as both a sensory gatekeeper (a critical ‘sensory switchboard’ often implicated in autism) and a pacemaker for cortical oscillations^11^. Recent experimental work has established a central role for the thalamus in the generation of generalized seizures, highlighting how altered thalamocortical circuit physiology—including bidirectional interactions between neurons and thalamic astrocytes—promotes the development of generalized spike-wave discharges^12^. In SYNGAP1 haploinsufficiency, synaptic plasticity deficits likely disrupt these circuits, manifesting electrically as generalized discharges (clinically presenting most prominently as atypical absences).

This thalamic dysfunction extends directly into the behavioral domain. The thalamus acts as the brain’s “sensory switchboard,” and recent translational ASD models demonstrate that hyperexcitability within the thalamic reticular nucleus directly drives core autistic behaviors, including social deficits and repetitive actions, even independently of spontaneous seizure activity^13^. This aligns perfectly with our clinical findings that the qualitative thalamocortical circuitopathy—rather than quantitative seizure frequency—dictates the severity of the behavioral phenotype. Consequently, the severe caregiver-reported autistic behaviors in SYNGAP1-DEE can be understood as a failure in sensory filtering and processing driven by this specific network instability. The high prevalence of eyelid myoclonia (65.2%) reinforces this link, as this seizure type sits at the intersection of the visual system and motor control, potentially reflecting a hyperexcitability of the visual cortex and thalamic reticular nucleus often implicated in photosensitive DEEs.

### Comparison with Related Syndromes

The dominance of eyelid myoclonia and absences in our cohort draws clinical parallels to Jeavons syndrome (Epilepsy with Eyelid Myoclonia)^14,15^. However, unlike classic Jeavons syndrome, where cognition can remain intact, SYNGAP1-DEE presents with a pervasive neurodevelopmental impairment. Our findings suggest that while the seizure semiology mimics idiopathic generalized epilepsies, the underlying synaptic failure is more extensive, leading to the profound behavioral comorbidities observed. By distinguishing these specific seizure correlates from focal epilepsies, we provide evidence that SYNGAP1-DEE should be conceptualized as a system-wide synaptopathy affecting distinct integration centers (like the thalamus).

### Clinical Relevance

These findings have immediate clinical implications. The presence of atypical absence seizures could serve as a “red flag” for the development of more severe associated neuropsychiatric phenotypes.

Because we found that even low-frequency occurrences of these semiologies are associated with higher caregiver-reported autistic behavior scores, clinicians observing these seizure types in young children should initiate screening for autism spectrum disorders, even before social deficits become overt. Furthermore, therapeutic strategies in SYNGAP1-DEE might need to shift from broad-spectrum antiseizure medications to targeted treatments that stabilize thalamocortical oscillations, potentially ameliorating both the seizures and the sensory processing deficits associated with autism. Precision approaches like gene supplementation are currently under exploration to address this root pathology^16^.

### Limitations

#### Several limitations warrant consideration

First, the primary behavioral outcome was based on a single caregiver-reported Likert item rather than standardized clinician-administered instruments (e.g., ADOS-2). Although caregiver reports can capture everyday functioning, they are susceptible to subjectivity, differential interpretation, and reporting bias, and may not map one-to-one onto a formal ASD phenotype.

Second, this registry-based analysis is retrospective and cross-sectional. Accordingly, causal inference is not possible: we cannot determine whether specific seizure semiologies contribute to behavioral severity, whether behavioral severity influences seizure recognition/reporting, or whether both represent parallel manifestations of the underlying SYNGAP1-related circuit dysfunction. In addition, potential confounding by age, treatment history, and comorbidity burden cannot be fully excluded in a univariate framework.

Third, seizure semiology and peak (maximal historical) seizure frequency were caregiver-reported and therefore prone to misclassification and recall bias. Differentiating atypical absences from brief focal impaired awareness seizures, recognizing subtle epileptic spasms, or distinguishing epileptic from non-epileptic motor phenomena can be challenging without contemporaneous video-EEG confirmation. Consistent with prior cohorts, focal seizures were reported in a minority of participants (∼9%), supporting a broader phenotypic spectrum in SYNGAP1-DEE than purely generalized epilepsy models. Reports of “epileptic spasms” may, in some cases, reflect other seizure patterns (e.g., myoclonic clusters) or non-epileptic movements.

Fourth, the seizure frequency variable represents an ordinal peak-frequency rating and does not capture cumulative seizure burden, seizure duration, clustering, or longitudinal dynamics. The absence of an association between this ordinal metric and caregiver-reported autistic behaviors should therefore be interpreted as “no detectable association within the limits of this measurement approach,” rather than definitive evidence that seizure burden is irrelevant.

Finally, restricting the analytic cohort to participants who completed both the epilepsy questionnaire and the behavioral questionnaire may introduce selection bias toward more engaged families or those with more salient symptoms; the direction of this bias is uncertain.

## 6. Conclusion

Our findings suggest a paradigm shift in SYNGAP1-DEE, moving beyond the generalized view that “epilepsy disrupts development” toward a precision model where atypical absences could serve as an indicator for a specific thalamocortical network dysfunction that drives distinct neurobehavioral deficits like epilepsy and autism. The association between the atypical absence phenotype and caregiver-reported autistic behavior scores underscores the necessity of considering the underlying circuitopathy rather than merely suppressing general seizure burden.

## References

1. Holder JL, Hamdan FF, Michaud JL. SYNGAP1-Related Intellectual Disability. In: Adam MP, Bick S, Mirzaa GM, Pagon RA, Wallace SE, Amemiya A, Herausgeber. GeneReviews® [Internet]. Seattle (WA): University of Washington, Seattle; 1993 [zitiert 14. Februar 2026]. Verfügbar unter: http://www.ncbi.nlm.nih.gov/books/NBK537721/

2. Hamdan FF, Gauthier J, Spiegelman D, Noreau A, Yang Y, Pellerin S, u. a. Mutations in SYNGAP1 in autosomal nonsyndromic mental retardation. N Engl J Med. 5. Februar 2009; 360(6):599–605.

3. Clement JP, Aceti M, Creson TK, Ozkan ED, Shi Y, Reish NJ, u. a. Pathogenic SYNGAP1 mutations impair cognitive development by disrupting maturation of dendritic spine synapses. Cell. 9. November 2012; 151(4):709–23.

4. McTague A, Howell KB, Cross JH, Kurian MA, Scheffer IE. The genetic landscape of the epileptic encephalopathies of infancy and childhood. Lancet Neurol. März 2016; 15(3):304–16.

5. Scheffer IE, French J, Valente KD, Auvin S, Cross JH, Specchio N. Operational definition of developmental and epileptic encephalopathies to underpin the design of therapeutic trials. Epilepsia. April 2025; 66(4):1014–23.

6. von Stülpnagel C, van Baalen A, Borggraefe I, Eschermann K, Hartlieb T, Kiwull L, u. a. Network for Therapy in Rare Epilepsies (NETRE): Lessons From the Past 15 Years. Front Neurol. 2020; 11:622510.

7. EURAS: EUropean network for neurodevelopmental RASopathies | EURAS [Internet]. [zitiert 14. Februar 2026]. Verfügbar unter: https://euras-project.eu/

8. Harris PA, Swafford J, Serdoz ES, Eidenmuller J, Delacqua G, Jagtap V, u. a. MyCap: a flexible and configurable platform for mobilizing the participant voice. JAMIA Open. Juli 2022; 5(2):ooac047.

9. Scheffer IE, Berkovic S, Capovilla G, Connolly MB, French J, Guilhoto L, u. a. ILAE classification of the epilepsies: Position paper of the ILAE Commission for Classification and Terminology. Epilepsia. April 2017; 58(4):512–21.

10. R: The R Project for Statistical Computing [Internet]. [zitiert 14. Februar 2026]. Verfügbar unter: https://www.r-project.org/

11. Woodward ND, Giraldo-Chica M, Rogers B, Cascio CJ. Thalamocortical dysconnectivity in autism spectrum disorder: An analysis of the Autism Brain Imaging Data Exchange. Biol Psychiatry Cogn Neurosci Neuroimaging. Januar 2017; 2(1):76–84.

12. Lindquist BE, Timbie C, Voskobiynyk Y, Paz JT. Thalamocortical circuits in generalized epilepsy: Pathophysiologic mechanisms and therapeutic targets. Neurobiol Dis. 1. Juni 2023; 181:106094.

13. Jang S-S, Takahashi F, Huguenard JR. Reticular thalamic hyperexcitability drives autism spectrum disorder behaviors in the Cntnap2 model of autism. Sci Adv. 22. August 2025; 11(34):eadw4682.

14. Jeavons PM. Nosological problems of myoclonic epilepsies in childhood and adolescence. Dev Med Child Neurol. Februar 1977; 19(1):3–8.

15. Vlaskamp DRM, Shaw BJ, Burgess R, Mei D, Montomoli M, Xie H, u. a. SYNGAP1 encephalopathy: A distinctive generalized developmental and epileptic encephalopathy. Neurology. 8. Januar 2019; 92(2):e96–107.

16. Quinlan MA, Guo R, Clark AG, Luber EM, Christian RJ, Martinez RA, u. a. AAV delivery of full-length SYNGAP1 rescues epileptic and behavioral phenotypes in a mouse model of SYNGAP1-related disorders. Mol Ther. 3. Dezember 2025; 33(12):6398–414.

